# Additive associations of variants at the expression quantitative trait loci of *TFR2* and *EPO* in a linkage disequilibrium block with hypochromic erythropoiesis

**DOI:** 10.1101/2025.09.12.25335626

**Authors:** Taku Nakai, Kinuko Ohneda, Takanori Hidaka, Katsuyuki Sasaki, Shin-ichi Fujimaki, Tohru Fujiwara, Hideo Harigae, Masayuki Yamamoto, Fumiki Katsuoka, Norio Suzuki

## Abstract

Reciprocal interactions between iron metabolism and erythropoiesis are well established in patients and animal models under stress. To elucidate this interplay in healthy individuals, we conducted a genome-wide association study (GWAS) involving 52,822 individuals of Japanese ancestry, and integrated the results with plasma biochemical parameters related to iron metabolism. We identified 30 genetic variants associated with erythrocyte traits, four of which also influenced serum iron concentrations and/or transferrin saturation within normal physiological ranges. These findings indicate a close interaction between erythropoiesis and iron metabolism, even in the absence of disease. Among the identified variants, we focused on rs2075672, an expression quantitative trait locus (eQTL) for *TFR2* (transferrin receptor 2). This variant resides within a linkage disequilibrium (LD) block that also includes rs1617640, a known causal variant at the *EPO* (erythropoietin) eQTL for increased erythrocyte counts. Individuals carrying this multiple causal variant-containing LD (mcv-LD) block exhibited mild hypochromic erythropoiesis. Our analyses revealed that the minor allele of rs2075672 is correlated with decreases in serum iron and erythrocytic hemoglobin concentrations, whereas the minor allele of rs1617640 appeared to primarily drive an increase in the erythrocyte count. Longitudinal follow-up analyses reproduced the original data and confirmed the persistence of hypochromic erythropoiesis in mcv-LD block carriers for at least five years. These data highlight the existence of mcv-LD blocks, carriers of which present unique phenotypes characterized by combined traits associated with each causal variant within the block.

**Graphical abstract:** 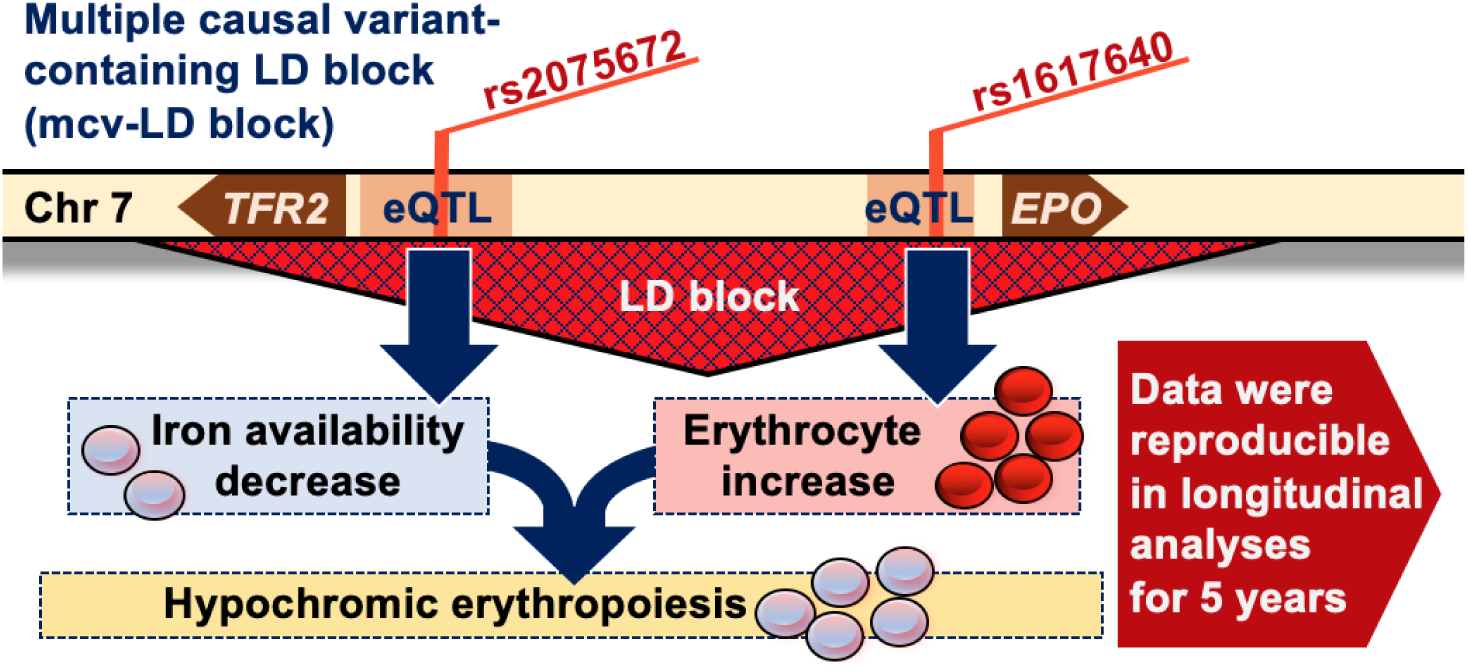

## Introduction

Iron, an essential metal for organisms, is most abundant in red blood cells (erythrocytes) as a component of the oxygen-carrying molecule hemoglobin among mammalian tissues. Owing to its highly toxic nature, which can lead to free radical formation, systemic mechanisms strictly regulate iron metabolism in a semi-closed manner [1]. Indeed, numerous studies have shown that erythropoiesis and iron metabolism are closely linked to maintaining oxygen delivery to all organs under stressed conditions in patients and animal models [2]. However, it is unclear whether iron metabolism and erythropoiesis are reciprocally fine-tuned within the healthy range in nondiseased individuals.

Using mouse models, we and others have shown that the erythroid growth factor erythropoietin (EPO) plays a critical role in promoting both erythrocyte increase and iron use for hemoglobin synthesis [3–5]. The *EPO* gene is expressed by kidney interstitial fibroblasts, referred to as renal EPO-producing (REP) cells, in a hypoxia (low oxygen supply)-inducible manner [6–8]. EPO from the kidney stimulates proliferation, anti-apoptosis and heme synthesis in erythroid precursors by binding to its specific receptor (EPOR) [9,10]. Additionally, the EPO-EPOR signal induces the secretion of erythroferrone, which suppresses the hepatic production of hepcidin. Hepcidin, which is responsible for iron storage and iron uptake, inhibits the release of iron from cells by blocking the iron exporter ferroportin [11–13].

After iron is released into the bloodstream, it is caged in transferrin and captured by cells expressing transferrin receptors (TFRs). There are two isoforms of TFRs, TFR1 and TFR2 [14]. TFR1 expression is widely observed on the surfaces of iron-incorporating cells and is strictly regulated to control intracellular iron levels [15]. TFR2 is principally expressed on the cell surfaces of erythroid precursors and hepatocytes to monitor blood iron levels by binding to transferrin [16,17]. In erythroid cells, TFR2 associates with EPOR to increase EPO-EPOR signaling, followed by iron use for hemoglobin synthesis when blood iron concentrations are high [18]. In hepatocytes, TFR2 also serves as an iron sensor, and transferrin-TFR2 binding increases the expression and secretion of hepcidin to reduce serum iron levels [19].

Many genetic variants have been found to affect erythropoiesis directly or indirectly via altering iron metabolism [20–23]. There are single-nucleotide variants (SNVs) that modulate the transcriptional levels of the *EPO* gene in the proximal promoter region [24,25], of which rs1617640 is known as an expression quantitative trait locus (eQTL) that affects serum EPO levels and erythropoiesis [26,27]. In addition to the proximal promoter region, relatively common variants are located in the distal upstream and proximal downstream regions of the *EPO* gene, which we previously reported as the renal and hepatic *cis*-regulatory elements of the gene, respectively [28,29]. The *TFR2* gene 100 kb upstream of the *EPO* gene on chromosome 7 harbors rare variants that lead to amino acid substitutions such as M172K and C282Y as causative mutations of hereditary hemochromatosis type 3 [30,31]. Moreover, eQTLs of the *TFR2* gene have been identified from common variants in the promoter and intron regions, which are subtly associated with iron metabolism disorders [32–34].

Recently, genetic variants associated with diseases and traits have been identified through genome-wide association studies (GWASs) based on large-scale population cohorts [35]. The Tohoku Medical Megabank (TMM) project represents the largest population cohort study, comprising over 150,000 residents from the Miyagi and Iwate prefectures in Japan. The project has established an integrated biobank that includes detailed physiological and genetic data from the participants [36–38]. An SNV array dataset was collected from nearly all participants using the Affymetrix Axiom Japonica Array, a population-specific SNV array developed exclusively for the Japanese population. This array facilitates the high-precision identification of ethnicity-specific variants associated with various phenotypes [39]. These cohort data provide a valuable resource for investigating variations in multi-omics data within a well-characterized Japanese biobank [40,41].

We conducted a GWAS on erythrocyte traits using the biobank of the TMM project to elucidate the complex relationship between erythropoiesis and iron metabolism in non-diseased individuals. We found that erythropoiesis and iron metabolism mutually influence each other, even within the normal range, in individuals carrying genetic variants previously established as causal variants associated with erythrocyte and/or iron metabolism traits. Additionally, this GWAS revealed that two SNVs located at the eQTLs of the *TFR2* and *EPO* genes, which reside within the same linkage disequilibrium (LD) block, are additively associated with hypochromic erythropoiesis. While GWASs often face challenges in identifying multiple causal variants that are co-inherited within the same LD block [42], we successfully demonstrated that the co-inherited SNVs in the *TFR2* and *EPO* genes within an LD block are separately linked to decreased iron availability and increased erythrocyte numbers, respectively. Consequently, this LD block is recognized as a novel “multiple causal variant-containing LD (mcv-LD) block,” where two co-inherited causal variants additively impact a unique erythropoietic trait. Remarkably, a 5-year longitudinal follow-up study reproduced the original data and confirmed the persistent presence of hypochromic erythropoiesis in carriers of this mcv-LD block.

## Materials and methods

### Legal and ethical compliance

This study was conducted in accordance with the Ethical Guidelines for Medical and Health Research Involving Human Subjects issued by the Ministry of Education, Culture, Sports, Science and Technology, the Ministry of Health, Labor and Welfare, and the Ministry of Economy, Trade, and Industry. This study was approved by the Research Ethics Review Board of Tohoku University (approval number: 2023-4-122).

### Collection of red blood cells and iron metabolism parameters

All blood samples were collected from participants of the TMM project [36–38]. TMM project established two prospective cohort studies: the “TMM Community-Based Cohort Study”, which involved 66,034 participants, and the “TMM Birth and Three-Generation Cohort Study”, which involved 73,529 participants (Figure 1) [43]. Red blood cell counts (RBC), hematocrit levels (Hct), and hemoglobin concentrations (Hb) were measured directly in the laboratory of LSI Medience (Tokyo), after which the mean corpuscular volume (MCV), mean corpuscular Hb content (MCH), and mean corpuscular Hb concentration (MCHC) were calculated (Table 1). For 2,095 participants in the TMM Community-Based Cohort Study cohort, the serum iron level (Fe), serum ferritin level (Frn), total iron-binding capacity (TIBC), unsaturated iron-binding capacity (UIBC), and transferrin saturation (TSAT) were also measured by Division of Laboratory Medicine, Tohoku University Hospital (Table 1). The 5-year longitudinal follow-up analysis was conducted with 11,438 participants in the TMM Community-Based Cohort Study in the fifth year following the initial analysis.

**Figure 1.**
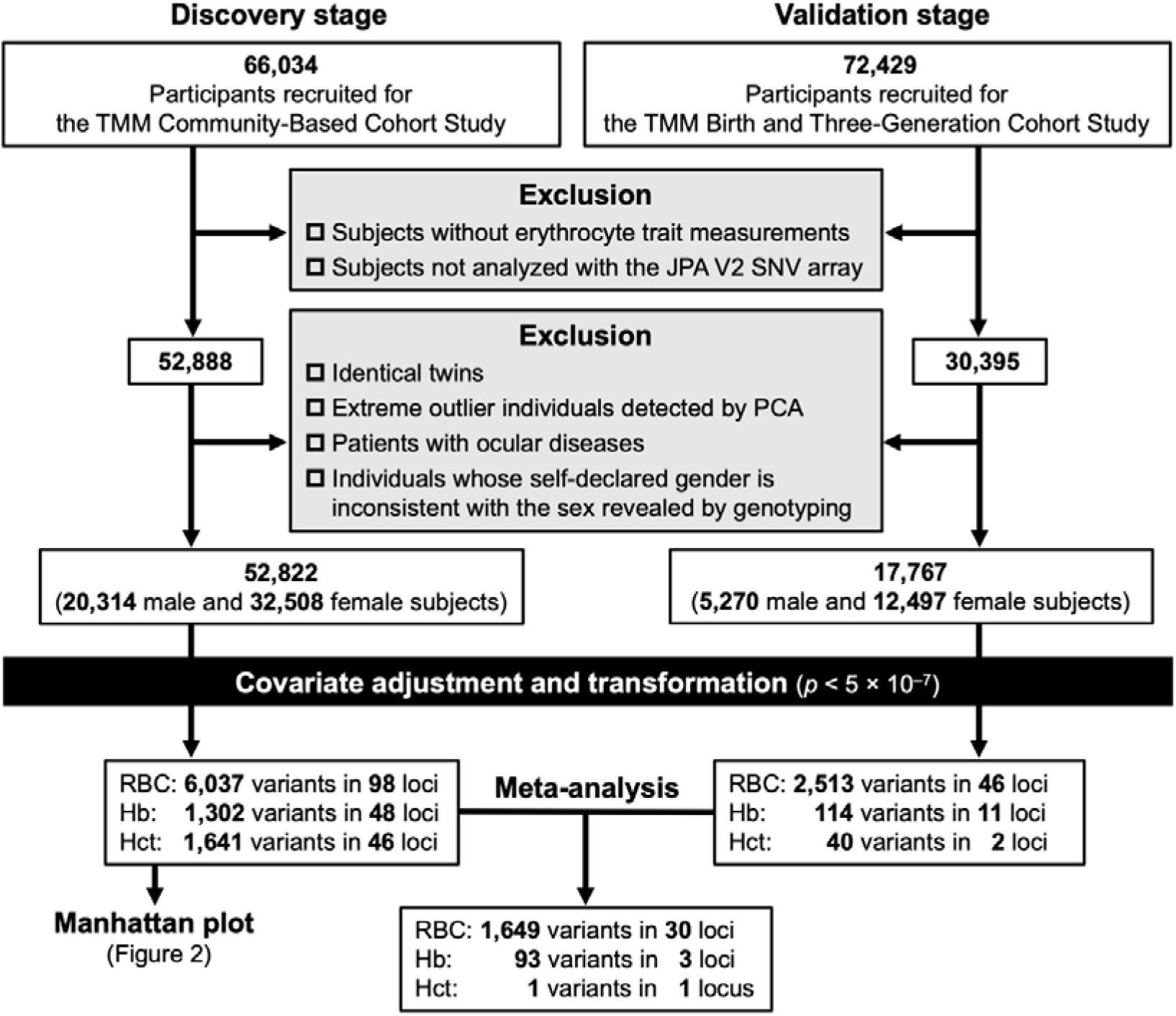
A study design flowchart for the current GWAS for erythrocyte traits. Data from the “TMM Community-Based Cohort Study” and the “TMM Birth and Three-Generation Cohort Study” were analyzed in the discovery and validation stages, respectively. PCA, principal component analysis.

**Table 1.**
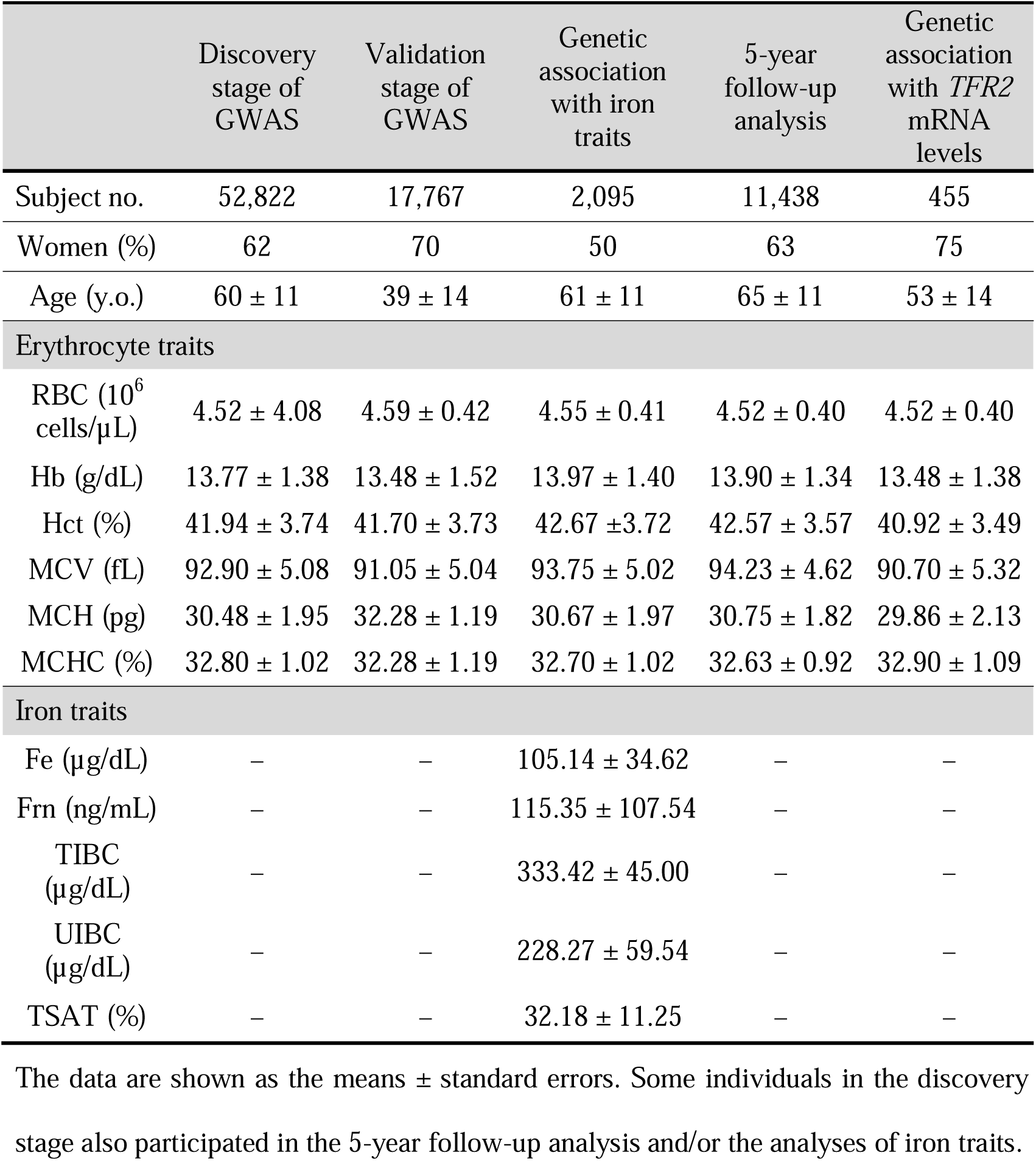
Baseline characteristics of all the subjects in each analysis of this study.

### Genotyping and quality control

Genotyping was performed using the Affymetrix Axiom Japonica Array V2 (JPA V2), a population-specific SNV array developed exclusively for the Japanese population, which consists of 654,246 tag variants [39]. Using the tag variants, imputation for untyped sites was carried out with a reference genome to increase the number of variants. For quality control, we excluded plates with an average call rate < 0.95 and removed samples with a DishQC metric < 0.82 before batch genotyping. We then applied a variant quality control step to each batch to exclude variants with *p* < 1.0 × 10^-5^ by a Hardy□Weinberg equilibrium test and a minor allele frequency < 0.01. We merged the imputed genotype datasets using QCTOOL (v2.0.4).

### GWAS

We conducted a linear mixed model analysis using BOLT-LMM v2.3.2 to evaluate the genetic effects of variants on 3 erythrocyte traits, RBC, Hct and Hb. To address potential biases from population stratification, familial connections and cryptic relatedness, the genetic correlation matrix was incorporated into the linear mixed model. Age and sex were included as covariates in the adjustment process for the erythrocyte traits. For the GWAS of each trait, both directly genotyped data and imputed genotype data were analyzed using BOLT-LMM. Associated variants identified in the discovery stage of the GWAS with the TMM Community-Based Cohort Study data in the discovery stage were validated using the TMM Birth and Three-Generation Cohort Study data in the validation stage. The variants that validated their significant associations with the erythrocyte traits were subsequently tested by a fixed-effect inverse-variance meta-analysis using effect estimates and standard errors from the discovery and validation stages [44]. A single locus containing multiple variants was defined as a 1.0-Mb genomic region (± 500 kb window from a variant). The nearest gene to a variant was designated as the putative gene linked to the variant.

### Transcriptome analysis

Transcriptome analysis was previously conducted using peripheral blood samples from 597 participants in the TMM Community-Based Cohort Study [45]. The data are available in the “Japanese Multi-omics Reference Panel (jMorp)” database (https://jmorp.megabank.tohoku.ac.jp)

### Statistical analyses

GWAS and subsequent analyses were conducted via R software (R Core Team) and PLINK2. For data preparation, PLINK2 was used for the conversion of BGEN-format files to BED-format files. We filtered variants with a minor allele frequency of at least 0.05 to ensure that only common variants were considered. We subsequently used the KING cutoff method to remove closely related individuals (cutoff = 0.0625) and reduced collinearity among variants by LD pruning with a window size of 50 variants, a step size of 5, and an r^2^ threshold of 0.2. With the above methods, we extracted independent variants and conducted principal component analyses (PCAs) to calculate the top 10 principal components, which were used to adjust for population stratification. After the cleaned genotype data were obtained, association analyses for each phenotype were performed using linear models. To control for population stratification, covariates such as age, sex, and the top 10 principal components were adjusted. Hierarchical clustering analyses of the variants were performed on the basis of patterns of significant fluctuation from the linear models to group variants with similar effects on erythrocyte traits and iron-related parameters.

## Results

### GWAS for erythrocyte traits in individuals of Japanese ancestry

The associations of genetic variations with 3 erythrocyte traits (RBC, Hb, and Hct) in 52,822 individuals were analyzed in the discovery stage of this GWAS using a dataset from the TMM Community-Based Cohort Study of individuals of Japanese ancestry (Table 1 and Figure 1) [38]. The results from the discovery stage were validated with the dataset from the TMM Birth and Three-Generation Cohort Study of individuals of Japanese ancestry in the meta-analysis stage, and a total of 1,743 variants in 30 loci were identified to be significantly associated with any of the erythrocyte traits (Table 2 and Figure 2). Associations of all variants identified in this GWAS with erythropoiesis have already been established across ancestries including Japanese [20–23].

**Figure 2.**
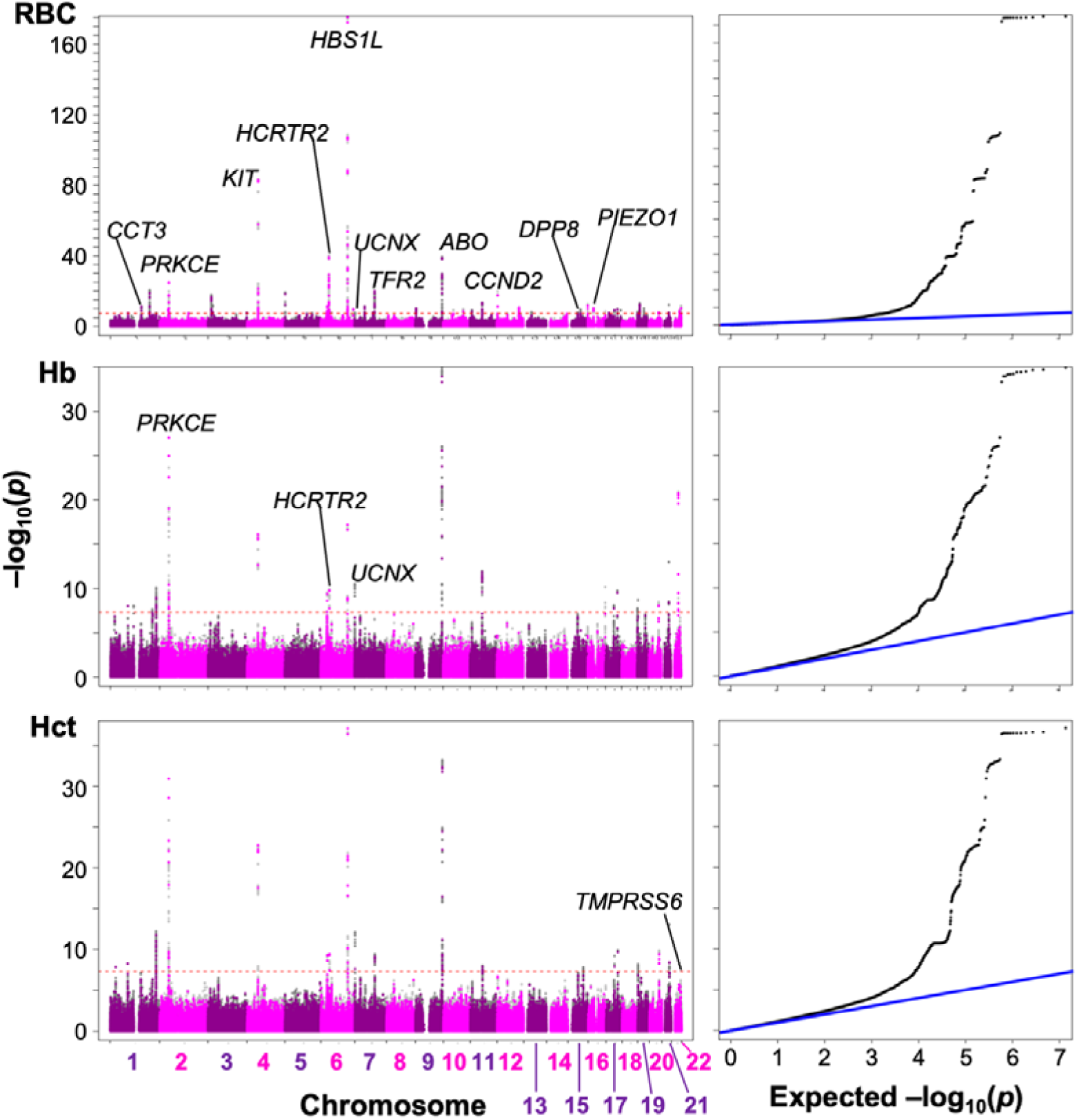
Manhattan plots (left) and quantile□quantile plots (right) from the GWAS for erythrocyte traits in the discovery stage. The vertical axes represent *p* values for the associations between genetic variants and each erythrocyte trait (RBC, Hb and Hct). The red dotted lines in the left panels indicate *p* = 1.0×10^−7^. Quantilel□quantile plots of observed versus expected *p* values under the null hypothesis are shown with the null expectation (blue lines) in the right panels.

**Table 2.**
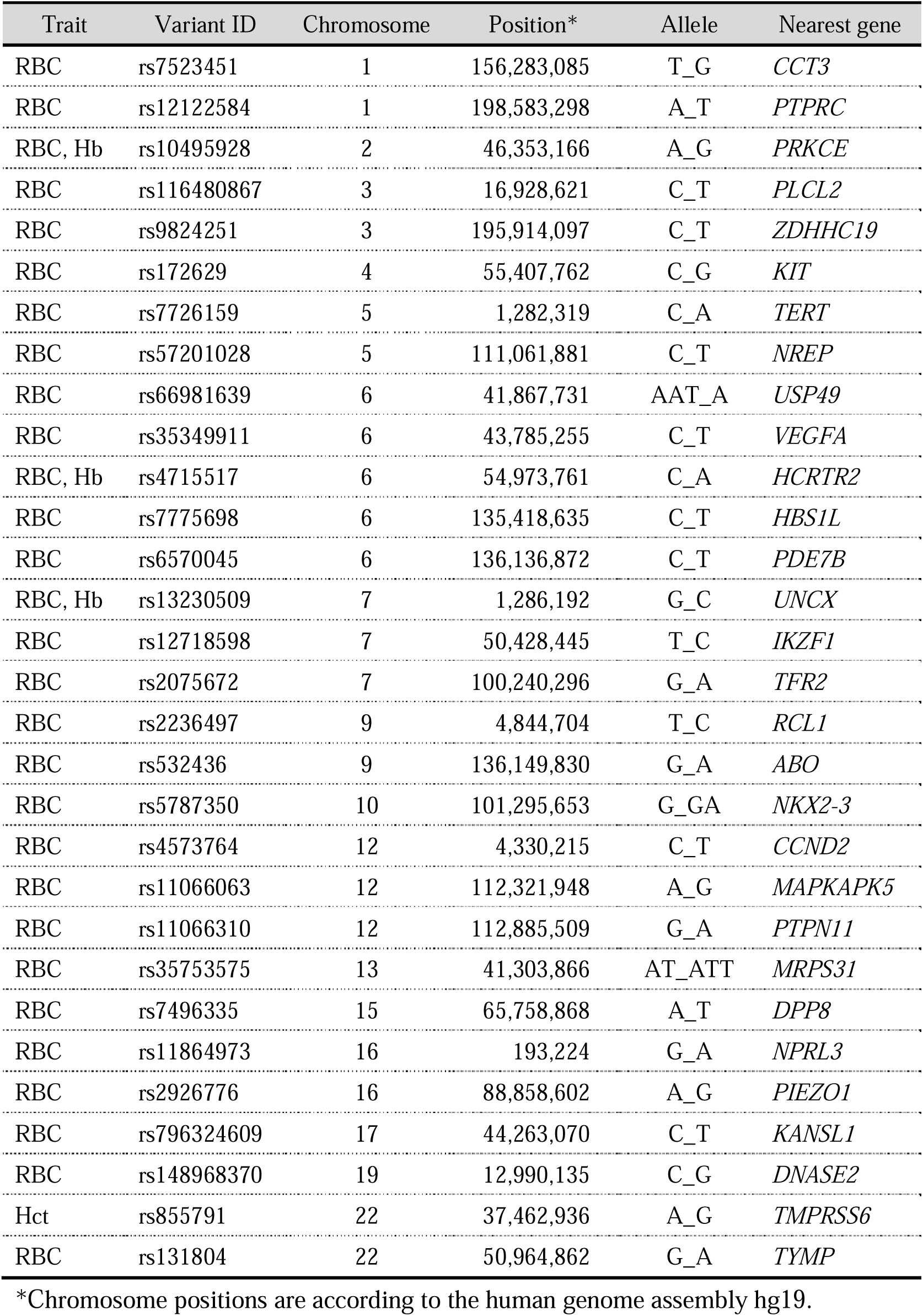
Genetic variants identified in this GWAS for erythrocyte traits.

On the basis of the minor allele signatures for RBC, the 30 identified variants were divided into high and low RBC groups. In the high RBC group, minor alleles of the 13 variants were associated with an increase in RBC, and 3 of which were also associated with increases in Hb and/or Hct (Figure 3). The other 17 minor alleles were classified into the low RBC group and presented decreased RBC, Hb, and/or Hct. In the low RBC group, the minor rs855791carriers uniquely exhibited increased Hb and Hct (Figure 3). Because rs855791 is a missense variant of the *TMPRSS6* gene encoding matriptase-2, an inhibitor of hepcidin production [46], its minor allele was thought to indirectly result in macrocytic-hyperchromic erythropoiesis with decreased RBC by affecting iron metabolism.

**Figure 3.**
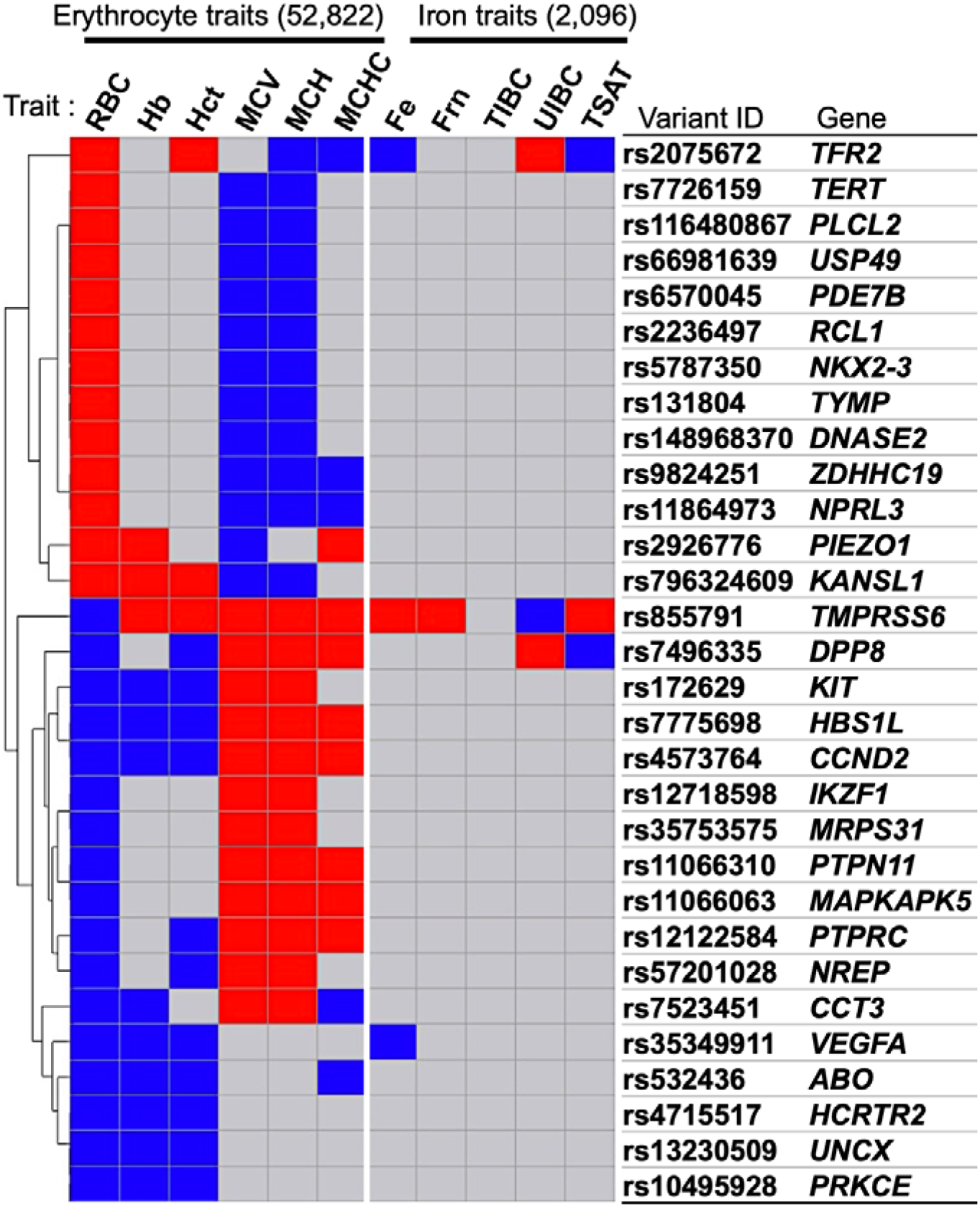
Genetic variants significantly associated with traits of erythrocytes and iron utilization. Genetic variants associated with increases (red) and decreases (blue) in erythrocyte traits (RBC, Hb, MCV, MCH and MCHC; n = 52,822) and iron utilization traits (Fe, Frn, TIBC, UIBC and TSAT; n = 2,095) compared with their mean values are shown in order of hierarchical clustering (tournament bracket). The nearest protein-coding genes from each variant are also listed.

Using data from the meta-analysis stage of this GWAS for RBC, Hb and Hct, the relationships between changes in these 3 erythrocyte traits and the other erythroid parameters, MCV, MCH, and MCHC, were examined. The results demonstrated that the minor alleles in the high RBC group, except rs2926776 (*PIEZO1*), are associated with increased production of abnormal erythrocytes to compensate for the decreased Hb in each erythrocyte (MCH), which is probably caused by their small size (MCV) and/or low Hb (MCHC, Figure 3). Since genetic variants of the *PIEZO1* gene, which encodes an ion channel, are linked to hereditary stomatocytosis through osmotic changes in erythrocytes [47], the exceptional MCHC increase in the high RBC group was probably caused by a reduction in the MCV of the individuals carrying the minor rs2926776 allele.

The minor allele signatures of the low RBC group were divided into 2 groups. MCV, MCH or MCHC were likely increased to compensate for the reduced number of erythrocytes in one subgroup, whereas they were unaffected or decreased in the other subgroup (Figure 3). In the latter subgroup, the minor allele of rs7523451 (*CCT3*) was exclusively related to macrocytic hypochromic erythropoiesis (Figure 3). The intronic variant may alter the expression levels of CCT3, which is involved in the recycling of TFRs [48], resulting in the modulation of iron intake into erythroid cells.

### Effects of genetic variants associated with erythrocyte traits on iron availability for hemoglobin synthesis

The traits of iron availability for hemoglobin synthesis (Fe, Frn, and transferrin states) were measured in 2,095 individuals who participated in the discovery stage (Table 2). Owing to the small sample size compared with that of the original GWAS for erythrocyte traits, only 4 of the 30 genetic variants were significantly associated with the iron traits (Figure 3). Among the 4 variants, only rs855791, which is a missense variant of the gene encoding the hepcidin suppressor matriptase-2 as described above, was associated with increases in both the serum iron (Fe) and iron storage (Frn) levels (Figure 3) [34]. Thus, reduced hepcidin activity in minor rs855791 carriers may provoke dietary iron intake and relative iron overload conditions, followed by increased production of hemoglobin [46].

rs7496335 is an intronic variant of the gene encoding dipeptidyl peptidase VIII (DPP8), which is involved in iron metabolism [34]. Our data suggested that the minor rs7496335 allele promotes iron use for hemoglobin synthesis in erythroid cells along with decreased TSAT followed by the production of macrocytic hyperchromic erythrocytes (Figure 3). rs35349911 is located proximal to the gene encoding vascular endothelial cell growth factor-A (VEGF-A) [49], suggesting that VEGF-A-mediated alterations in vascular permeability and/or fragility may be related to decreased serum Fe levels in minor rs7496335 carriers (Figure 3). rs2075672 (*TFR2*) is a known variant associated with iron homeostasis [34], and its minor allele may influence compensatory induction of erythrocyte production for reduced hemoglobin synthesis in each erythrocyte owing to a lack of serum iron (Figure 3). Collectively, this GWAS for erythrocyte and iron traits revealed that erythropoiesis, hemoglobin synthesis, iron availability and iron storage are reciprocally controlled within the normal range to maintain oxygen delivery even under non-diseased conditions.

### The *TFR2-EPO* LD block contains two major variants associated with iron and erythrocyte traits

Since TFR2 and TMPRSS6 are positive and negative regulators of hepcidin production, respectively [50,51], our genome-wide analyses suggested that genetic alterations in iron metabolism influence erythropoietic regulation to maintain homeostasis of the oxygen supply in non-diseased humans (Figure 3). rs855791 (*TMPRSS6*) has been well characterized as a causal missense variant, the minor allele of which suppresses hepatic hepcidin production with increases in serum iron concentrations and transferrin saturation [52]. However, little is known about rs2075672 (*TFR2*), except its genomic location in a 300-kb LD block between the *TFR2* and *EPO* genes at 100,240,296 of chromosome 7 (Table 2 and Figure 4) [33].

**Figure 4.**
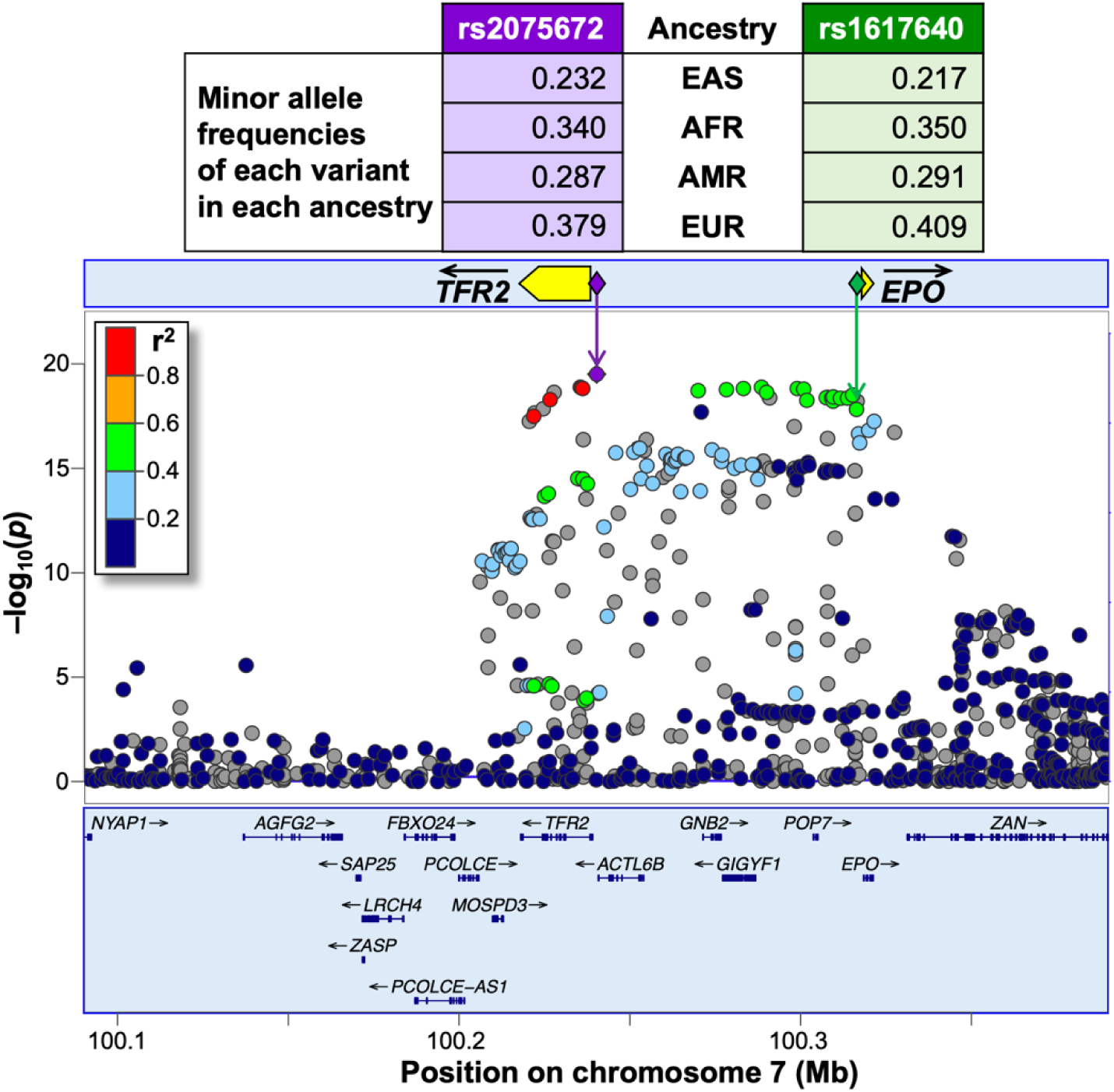
Associations of erythrocyte counts with genetic variants between *TFR2* and *EPO*. Zoom plots of the *p* values for the associations of RBC counts with genetic variants in chromosome 7q22.1. The squared correlation values (r^2^) of the linkage disequilibrium between rs2075672 (purple diamond) and each variant (circles) are shown with 5 grades by color. The gray dots indicate variants whose r^2^ values were not determined. The minor allele frequencies of rs2075672 and rs1617640 (green diamonds) in each ancestry of inhabitants in East Asia, Africa, America and Europe (EAS, AFR, AMR, and EUR, respectively) are also shown in the table on the basis of the human genome assembly hg19.

On the basis of the human genome assembly hg19, the minor rs2075672 allele (G for the major allele is replaced with A) in the *TFR2-EPO* LD block, which contains numerous RBC-associated variants, is present not only in individuals of East Asian ancestry but also in other individuals at frequencies of 23–38% (Figure 4). Among the variants in the *TFR2-EPO* LD block, rs1617640 has been characterized the most, and the causality of the minor rs1617640 allele (A for the major allele is replaced with C) for RBC and Hb increases has been reported [24,26]. The minor allele frequencies of rs1617640 and rs2075672 are comparable within each ancestry examined in hg19 (Figure 4), suggesting that these variants are contained in an identical LD block across ancestries. In fact, our data used for this discovery stage demonstrated that 14,408 (27.3%) and 15,290 (28.9%) of 52,822 individuals carried the minor rs2075672 and rs1617640 alleles, respectively, with 84.1% of the minor rs2075672 carriers also possessing minor rs1617640 (Table 3).

**Table 3.**
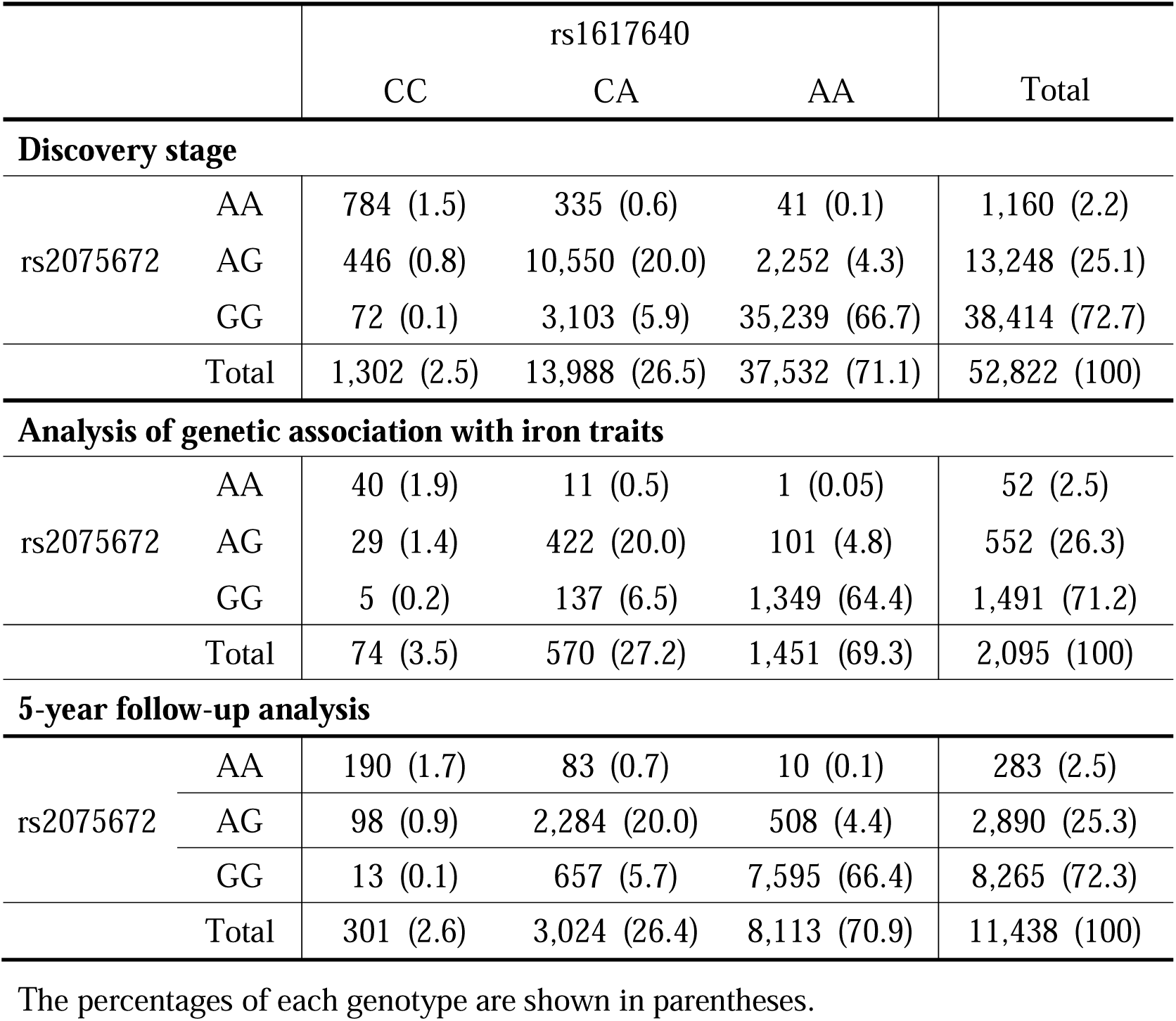
Numbers of participants carrying rs2075672 and/or rs1617640 in this GWAS.

There are numerous variants closely associated with RBC in the *TFR2-EPO* LD block other than rs2075672 and rs1617640. These variants are likely divided into 2 groups by the *ACTL6B* gene locus, each of which is represented by rs2075672 and rs1617640, respectively (Figure 4). The limited distribution of the variants belonging to the rs2075672 group around the *TFR2* gene predicted that *TFR2* is the functional target of these variants, among which rs2075672 has the lowest *p* value.

For the rs1617640 group, rs1617640 associated with an RBC increase has been exclusively verified [24, 26]. There is no evidence that the genes located between the *TFR2* and *EPO* genes (*ACTL6B*, *GNB2*, *GIGYF1* and *POP7*) are related to iron metabolism or erythropoiesis. These findings suggest that hypochromic erythropoiesis in the minor rs2075672 carriers may be associated with both or one of the two variants in the *TFR2-EPO* LD block.

### The combined association of two variants in the *TFR2*-*EPO* LD block with hypochromic erythropoiesis persists for at least 5 years

More than 80% of the minor rs2075672 carriers also bore the minor rs1617640 allele (Table 3). We then examined whether the hypochromic erythropoiesis observed in the minor rs2075672 carriers (Figure 3) was specifically related to rs2075672 or caused by the combined effects of multiple variants in the LD block. The associations of rs2075672 and rs1617640 with erythrocyte traits were separately analyzed using the data from the discovery stage.

The *p* values for the associations revealed that the decrease in the serum iron concentration was significantly affected by rs2075672 but not by rs1617640 (Figure 5A). The RBC and Hct levels were influenced by both variants, whereas the impacts of rs1617640 were greater than those of rs2075672. An increase in Hb was exclusively associated with the minor rs1617640 allele but not the minor rs2075672 allele. Data from the analyses of individuals homozygously carrying the minor rs2075672 and/or rs1617640 alleles suggested that the iron reduction and RBC increase, which were discovered with the minor rs2075672 carriers in this GWAS, were independently affected by the variants in the *TFR2-EPO* LD block; the iron reduction is exclusively associated with rs2075672; and the RBC increase is associated predominantly with rs1617640.

**Figure 5.**
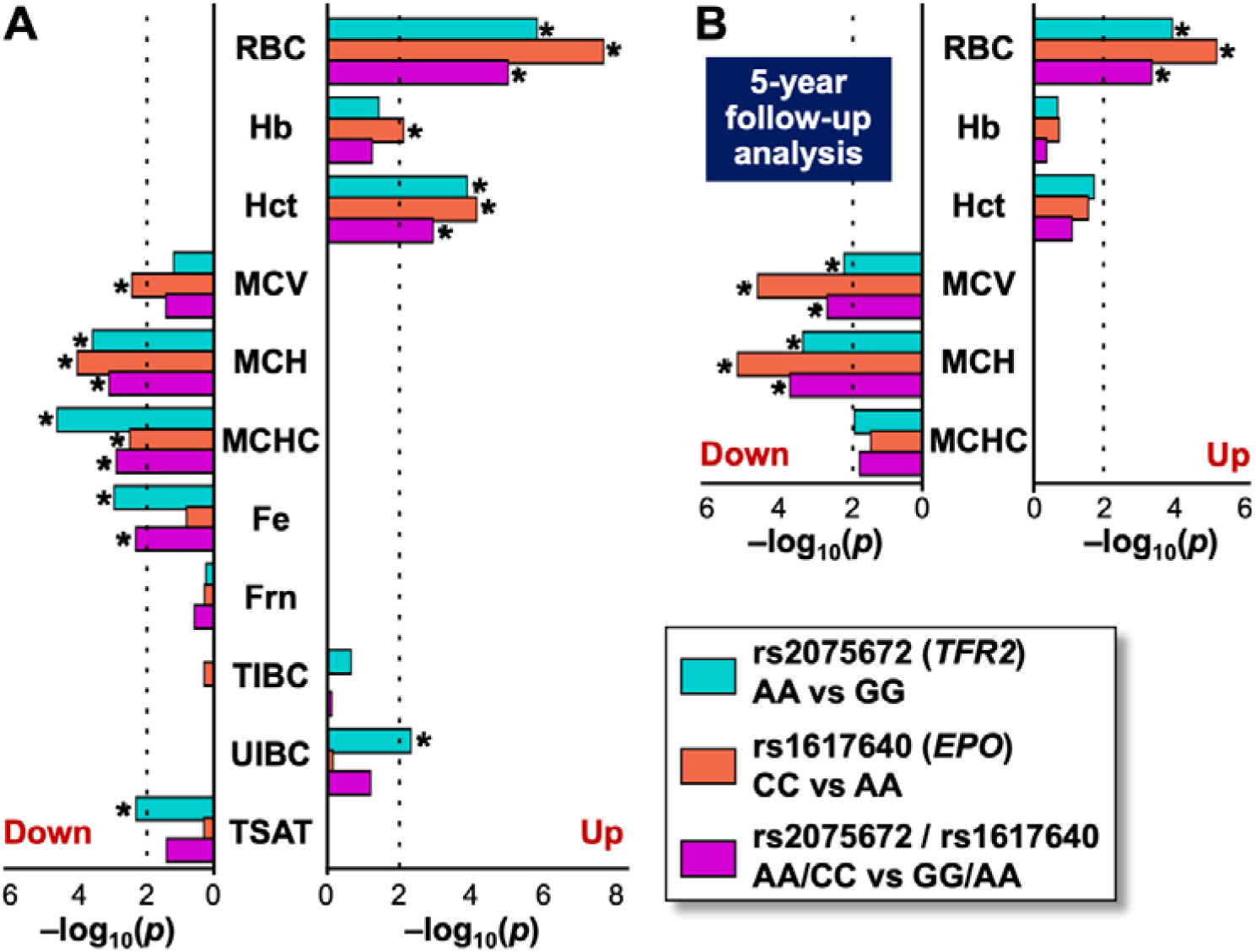
Effects of rs2075672 and/or rs1617640 on erythrocyte and iron utilization remain for 5 years in each individual. *p* values for the associations of the genetic variants with each trait in the original meta-analysis (A) and the 5-year follow-up analysis (B) were evaluated using a linear regression model that adjusted for the first 10 principal components, age, and sex. Asterisks indicate *p* = 0.01 (dotted line). “Up” and “Down” indicate that the values in the groups homozygously carrying the minor alleles (A for rs20756723; C for rs1617640) were increased and decreased, respectively, compared with those in the groups homozygously carrying the major alleles.

Utilizing the advantages of the TMM project, which possesses large-scale longitudinal data from follow-up cohort analyses, we tested the reproducibility of our original findings regarding the association between the *TFR2-EPO* LD block and hypochromic erythropoiesis. The erythrocyte traits of 11,438 individuals in the discovery stage of this GWAS were re-evaluated five years after the initial analyses (Tables 1 and 3). The results robustly confirmed that the minor alleles of rs1617640 and rs2075672 are cooperatively associated with hypochromic erythropoiesis. Additionally, we observed a sustained persistence of hypochromic erythropoiesis in individuals carrying both minor alleles in the *TFR2-EPO* LD block for at least five years (Figure 5B). These results strongly underscore the reliability of our original results and highlight the significant advantages provided by the TMM project.

### rs2075672 affects the expression levels of the *TFR2* gene in blood cells

rs2075672 was estimated to be located in the *TFR2* gene regulatory region. Indeed, on the basis of the GTEx database, hepatic *TFR2* mRNA levels in the minor rs2075672 (A) carriers were significantly greater than those in non-carriers (Figure 6A). TFR2 increases hepcidin production in the liver, which decreases serum iron concentrations by inhibiting the release of stored iron from reticular cells and dietary iron from duodenal epithelial cells [17,19,51]. We thus hypothesized that the reduced serum iron levels followed by reduced erythroid hemoglobin synthesis in the minor rs2075672 carriers (Figure 3) could be caused by increased expression of hepatic *TFR2*.

**Figure 6.**
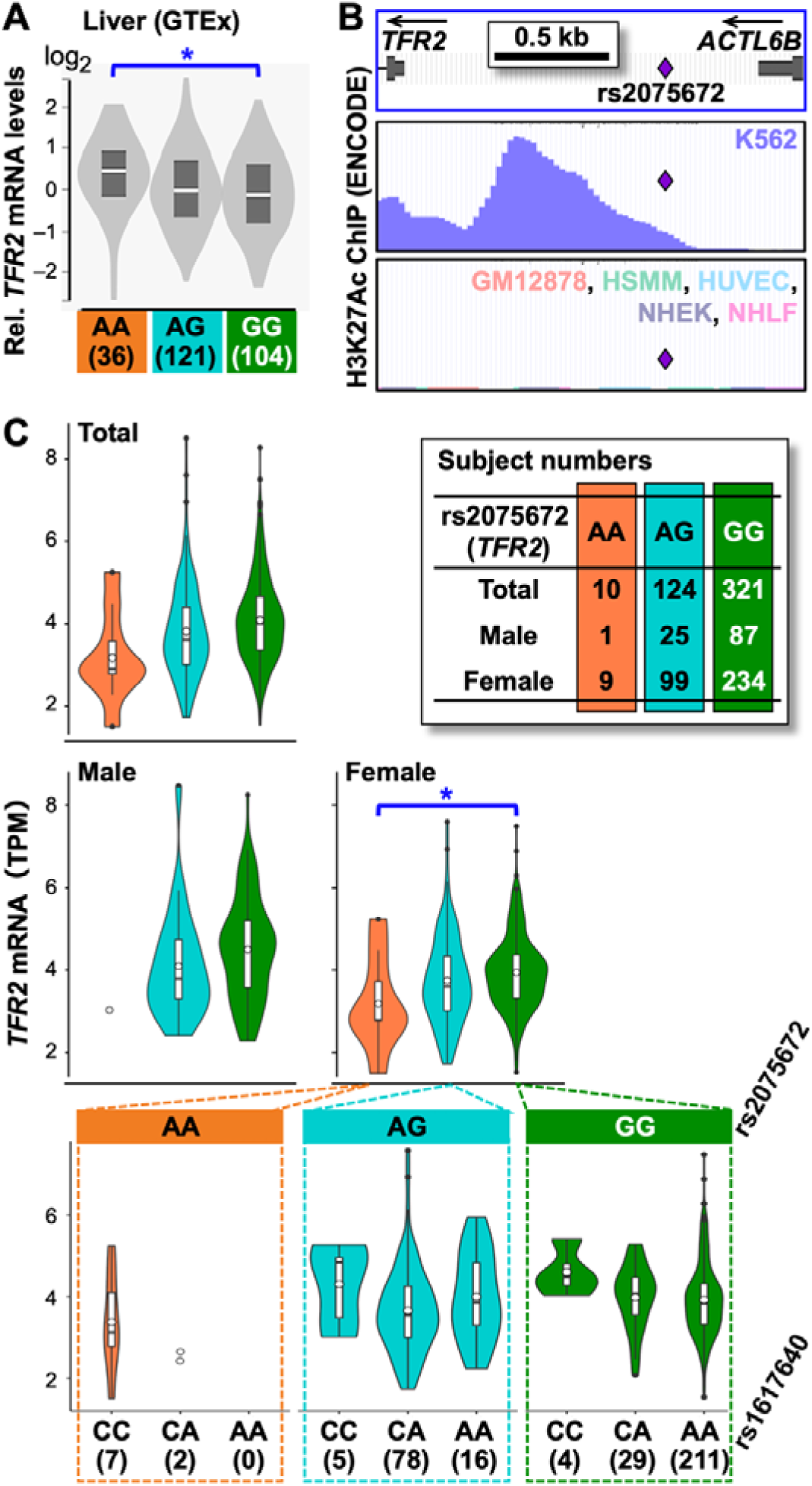
Different levels of *TFR2* mRNA expression in peripheral blood from individuals carrying the rs2075672 and/or rs1617640 variants. (A) Relative *TFR2* mRNA levels in the livers of individuals carrying rs2075672 variants. The data were obtained from GTEx. The numbers of individuals carrying each allele of rs2075672 are shown in parentheses. (B) Acetylation of histone H3 lysine 27 (H3K27Ac) in the *TFR2* gene promoter region of the K562 erythroid cell line (upper) was compared to that of the GM12878, HSMM, HUVEC, NHEK and NHLF cell lines. Chromatin immunoprecipitation (ChIP) data were obtained from the UCSC browser with the ENCODE database. (C) Violin plots showing *TFR2* mRNA levels (transcripts per million, TPM) from RNA-sequencing data of peripheral blood samples from total, male and female individuals carrying rs2075672 variants [44]. Data from female subjects carrying the rs2075672 and/or rs1617640 variants are also shown (lower graphs). The numbers of individuals carrying each allele of rs2075672 are shown in the upper right table, and those of rs1617640 are shown in the parentheses of the lower graphs. * *p* < 0.05 according to the Tukey-Kramer test.

In addition to hepatocytes, erythroid cells highly express *TFR2* to increase EPO-EPOR signaling for the proliferation and maturation of erythrocytes [18,53,54]. On the basis of the ENCODE database, high-level acetylation of histone H3 lysine 27 (H3K27Ac) in the *TFR2* gene promoter region was detected in the K562 erythroid cell line but not in the GM12878 (B-cell progenitor), HSMM (skeletal muscle myoblast), HUVEC (endothelial cell), NHEK (endothelial cell) or NHLF (lung fibroblast) cell lines (Figure 6B) [33]. Thus, rs2075672 in this epigenetically active region (Figure 6B) was estimated to be localized in the *TFR2* eQTL of erythroid lineage cells [55].

The genetic variant-associated expression of *TFR2* in erythroid cells was not included in the GTEx database. We then conducted an RNA-sequence analysis of peripheral blood with 455 participants in the discovery stage (Table 1) [44]. The data revealed that the minor rs2075672 allele tended to be associated with a reduction in *TFR2* mRNA levels in blood cells, although the reduction was not statistically significant (Figure 6C). The analysis of female *TFR2* mRNA levels, which were lower than those in males, revealed that *TFR2* mRNA levels in the blood cells of homozygous minor rs2075672 carriers were significantly lower than those in the blood cells of homozygous major rs2075672 carriers (Figure 6C). Because *TFR2* mRNA expression is undetectable or very low in hematopoietic cells except erythroid lineage cells (Figure 6B) [56], these data suggest that erythroid *TFR2* expression is attenuated by a single-nucleotide replacement at rs2075672.

Erythroid *TFR2* mRNA levels are influenced by erythropoietic activity and iron availability [17,19]. To exclude these indirect effects, the association of rs2075672 with erythroid *TFR2* expression was analyzed after stratifying the rs1617640 carriers, in which EPO levels were supposedly altered. The data suggested that the minor rs1617640 allele slightly enhances the expression of the *TFR2* gene neighboring the major rs2075672 allele (lower right panels in Figure 6C). However, *TFR2* mRNA levels were decreased, rather than increased, in peripheral blood cells homozygously carrying both minor alleles.

Our RNA-sequence analysis confirmed that the expression of the genes located in the *TFR2-EPO* LD block is undetectable (for *ACTL6B* and *EPO*) or unaffected by both variants in the *TFR2-EPO* LD block (for *GNB2*, *GIGYF1* and *POP7*) in peripheral blood cells. Collectively, the minor rs2075672 allele is associated with a decrease in erythroid *TFR2* expression. Moreover, it is considered that the RBC increase in individuals carrying both minor rs1617640 and rs2075672 alleles is associated with rs1617640 rather than rs2075672 because the loss of TFR2 in erythroid cells has been demonstrated to inactivate the proliferation and maturation of erythroblasts [18].

## Discussion

The current GWAS for erythrocyte traits in individuals of Japanese ancestry was conducted using the SNV array, JPA V2, developed exclusively for the Japanese population in the TMM project [39], and 30 variants significantly associated with erythrocyte traits were identified. Given the ancestral diversity in genetic variations, large-scale GWASs with non-European ancestries, such as this study, are crucial because most GWASs to date have been conducted using data from individuals of European ancestries [57]. All the variants identified in this study were already discovered by previous GWASs for erythrocyte traits [20–23], suggesting that the major erythrocyte-associated variants are common in European and East Asian ancestries. Although it is difficult to use experimental replicates in human subject analyses, this study successfully confirmed the solidity of the original GWAS data with reproducible data from the 5-year longitudinal follow-up analysis.

The data from this study indicated that blood erythrocyte concentrations, erythrocyte sizes, hemoglobin contents in erythrocytes, serum iron concentrations, iron storage (Frn), and transferrin states interact to maintain oxygen delivery homeostasis within the normal range under non-diseased conditions. The genetic variants identified in this GWAS affect one of these traits, leading to compensatory alterations in the other traits. Consequently, erythropoiesis and iron metabolism reciprocally influence in non-diseased humans, as we and others previously demonstrated in patients and animal models [3, 58–63].

Among the identified variants, we focused on rs2075672, which is located 1.0 kb upstream from the transcription start site of the *TFR2* gene, as its minor allele carriers uniquely exhibited an RBC increase and a decrease in serum iron. In the liver, the principal *TFR2*-expressing site, the GTEx database revealed that the minor rs2075672 allele is related to the upregulation of *TFR2* expression, indicating the eQTL localization of rs2075672. TFR2 in hepatocytes is involved in the production of hepcidin, a suppressor of iron release from iron-storing cells and duodenal epithelial cells into the blood [32,33, 51]. Thus, the decrease in serum iron concentrations in minor rs2075672 carriers is likely caused by increased hepcidin production. Additionally, the decrease in serum iron is thought to attenuate hemoglobin synthesis in each erythroid precursor, which has been observed in the minor rs2075672 carriers. Consequently, we propose that rs2075672 at the eQTL of *TFR2* is a causal variant associated with decreases in serum iron and MCH levels.

TFR2 is also expressed in erythroid cells and supports the EPO-EPOR signal to induce proliferation and maturation of the cells [17–19, 55]. rs2075672 is located in the erythroid-specific epigenetically active region of the *TFR2* gene promoter, and our data demonstrated that the minor rs2075672 allele is related to reduced *TFR2* expression in erythroid cells. The major rs2075672 allele (<u>A</u> of CGG<u>A</u>AG) is a part of a typical binding motif for ETS transcription factors, some of which play important roles in erythroid cells and hepatocytes [64], and the minor allele potentially makes the motif function invalid. Therefore, rs2075672 is considered directly associated with the expression levels of the *TFR2* gene in erythroid cells and hepatocytes.

These findings suggest that the decreases in MCH and MCHC levels observed in the minor rs2075672 carriers resulted from reduced iron availability for hemoglobin synthesis due to increased *TFR2* expression in hepatocytes and/or attenuated EPO-EPOR signaling due to decreased *TFR2* expression in erythroid cells. However, these changes in *TFR2* expression related to rs2075672 fail to explain the increase in RBC in minor rs2075672 carriers. We subsequently examined the contribution of rs1617640 to the RBC increase because rs1617640, a causal variant for RBC and Hb increases, is located at the *EPO* eQTL and is co-inherited with rs2075672 in an identical LD block [24]. As expected, the data in this study demonstrated that the minor rs1617640 allele is associated with RBC increase, regardless of serum iron level or rs2075672.

In summary, we concluded that the two co-inherited variants at the *TFR2* and *EPO* eQTLs are independently associated with decreased iron availability and increased erythrocyte numbers in normal ranges, respectively. Moreover, the *TFR2*-*EPO* LD block is one of the mcv-LD blocks associated with unique traits, such as hypochromic erythropoiesis, through independent contributions of each causal variant in the mcv-LD blocks. While recent high-precision and large-scale biobank resources have accelerated the detection of lead variants in GWASs, the identification of proximal causal variants remains challenging for current GWAS technology [42]. This study successfully identified co-inherited causal variants at the *TFR2* and *EPO* eQTLs for serum iron concentrations and RBC production, respectively. Additionally, these traits combinatorially contribute to hypochromic erythropoiesis, which is unique erythropoiesis characterized by the production of erythrocytes with low hemoglobin concentrations, in individuals carrying this mcv-LD block.

## Data Availability

All data produced in the present study are available upon reasonable request to the authors.

## Data and code availability

All data generated by this study are contained in the published article.

## Acknowledgements

We would like to thank the TMM cohort study participants, members of the TMM project (https://www.megabank.tohoku.ac.jp/english/a240901/) that is supported by grants from the Japan Agency for Medical Research and Development (AMED; JP17km0105001, JP21tm0124005, and JP21tm0424601) as well as the clinical laboratory of Tohoku University Hospital. This study was supported by the Ministry of Education, Culture, Sports, Science and Technology (MEXT; 23KK0137, 25H01437 to N.S., and 23K144500 to T.N.), the Frontier Research in Duo (FRiD) of Tohoku University Research Program, the Astellas Foundation for Research on Metabolic Disorders, the Naito Foundation, the Uehara Memorial Foundation, the Takeda Science Foundation (to N.S.) as well as the JKA Promotion Funds from KEIRIN RACE, the Mizuno Sports Promotion Foundation, and the Yamaha Motor Foundation for Sports (to T.N.). We also thank Mr. Daisuke Kubota and Ms. Atsuko Konuma (Tohoku University) for their technical assistance.

## Author contributions

Conceptualization, T.N. and N.S.; methodology, T.N., K.O., T.H. and F.K.; formal analysis, T.N., K.O., T.H., S-i.F, T.F. and H.H.; investigation, T.N. and T.H.; data curation, K.O. and N.S.; writing, T.N., K.O. and N.S.; supervision, M.Y., F.K. and N.S.; project administration, T.N. and N.S.; funding acquisition, T.N. M.Y. and N.S.

## Web resources

BOLT-LMM, https://alkesgroup.broadinstitute.org/BOLT-LMM/BOLT-LMM_manual.html

GTEx, https://www.gtexportal.org/home/

jMorp, https://jmorp.megabank.tohoku.ac.jp/

LocusZoom, https://my.locuszoom.org

PLINK2, https://www.cog-genomics.org/plink/2.0/

QCTOOL, https://www.chg.ox.ac.uk/∼gav/qctool_v2/

R, https://www.r-project.org/

UCSC Genome Browser, https://genome.ucsc.edu/

1000 Genomes Project, https://www.internationalgenome.org

